# Real-world clinical outcomes of treatment with casirivimab-imdevimab among patients with mild-to-moderate coronavirus disease 2019 during the Delta variant pandemic

**DOI:** 10.1101/2021.12.19.21268078

**Authors:** Yasuhito Suzuki, Yoko Shibata, Hiroyuki Minemura, Takefumi Nikaido, Yoshinori Tanino, Atsuro Fukuhara, Ryuzo Kanno, Hiroyuki Saito, Shuzo Suzuki, Taeko Ishii, Yayoi Inokoshi, Eiichiro Sando, Hirofumi Sakuma, Tatsuho Kobayashi, Hiroaki Kume, Masahiro Kamimoto, Hideko Aoki, Akira Takama, Takamichi Kamiyama, Masaru Nakayama, Kiyoshi Saito, Koichi Tanigawa, Masahiko Sato, Toshiyuki Kanbe, Norio Kanzaki, Teruhisa Azuma, Keiji Sakamoto, Yuichi Nakamura, Hiroshi Otani, Mitsuru Waragai, Shinsaku Maeda, Tokiya Ishida, Keishi Sugino, Yasuhiko Tsukada, Ryuki Yamada, Riko Sato, Takumi Omuna, Hikaru Tomita, Mikako Saito, Natsumi Watanabe, Mami Rikimaru, Takaya Kawamata, Takashi Umeda, Julia Morimoto, Ryuichi Togawa, Yuki Sato, Junpei Saito, Kenya Kanazawa, Ken Iseki

**Author notes:** **Correspondence:** All correspondence should be addressed to Yoko Shibata, MD, PhD., Address: Department of Pulmonary Medicine, Fukushima Medical University, School of Medicine, 1 Hikarigaoka, Fukushima City, Fukushima Prefecture, 960-1295, Japan. **Author contributions:** Conception and design: Yasuhito Suzuki and Yoko Shibata. Analysis and drafting the manuscript: Yasuhito Suzuki and Yoko Shibata. Data curation: all authors. Final approval of the manuscript: Yasuhito Suzuki and Yoko Shibata. Footnote page. Funding This study received no specific grant from any funding agency.

## Abstract

**Background:** Mutations of severe acute respiratory syndrome coronavirus 2 (SARS-CoV-2) may reduce the efficacy of neutralizing monoclonal antibody therapy against coronavirus disease 2019 (COVID-19). We here evaluated the efficacy of casirivimab-imdevimab in patients with mild-to-moderate COVID-19 during the Delta variant surge in Fukushima Prefecture, Japan.

**Methods:** We enrolled 949 patients with mild-to-moderate COVID-19 who were admitted to hospital between July 24, 2021 and September 30, 2021. Clinical deterioration after admission was compared between casirivimab-imdevimab users (n = 314) and non-users (n = 635).

**Results:** The casirivimab-imdevimab users were older (P < 0.0001), had higher body temperature (≥ 38°C) (P < 0.0001) and greater rates of history of cigarette smoking (P = 0.0068), hypertension (P = 0.0004), obesity (P < 0.0001), and dyslipidemia (P < 0.0001) than the non-users. Multivariate logistic regression analysis demonstrated that receiving casirivimab-imdevimab was an independent factor for preventing deterioration (odds ratio 0.448; 95% confidence interval 0.263–0.763; P = 0.0023). Furthermore, in 222 patients who were selected from each group after matching on the propensity score, deterioration was significantly lower among those receiving casirivimab-imdevimab compared to those not receiving casirivimab-imdevimab (7.66% vs 14.0%; p = 0.021).

**Conclusion:** This real-world study demonstrates that casirivimab-imdevimab contributes to the prevention of deterioration in COVID-19 patients after hospitalization during a Delta variant surge.

**Summary:** This real-world retrospective study demonstrates the contribution of treatment with casirivimab-imdevimab to the prevention of deterioration in patients with mild-to-moderate coronavirus disease 2019 (COVID-19) even during the Delta variant pandemic.

## 1. Introduction

Coronavirus disease 2019 (COVID-19), caused by severe acute respiratory syndrome coronavirus 2 (SARS-CoV-2), is continuing to spread around the world. Vaccines against SARS-CoV-2 have reduced infection and mortality and being widespread all over the world. Newly emerging variants have mutations in the spike protein of SARS-CoV-2 and show high infectivity [1]. Many neutralizing antibodies and vaccines against SARS-CoV-2 target this spike protein. Therefore, spike protein mutations may diminish their effectiveness [2]. Indeed, cases of the B.1.617.2 (Delta) variant, which possesses high transmissibility, spread throughout Japan from July to October 2021 [3].

Neutralizing monoclonal antibody therapy has been authorized by the United States Food and Drug Administration for treatment of high-risk patients with mild-to-moderate COVID-19 [4, 5]. Casirivimab-imdevimab is two recombinant human IgG1 monoclonal antibodies, known as REGN-COV2, that are used for the treatment of COVID-19 [5]. These antibodies are derived from humanized mice and the sera of patients recovering from COVID-19, and bind non-competitively to non-overlapping epitopes of the spike protein receptor-binding domain of SARS-CoV-2, thereby preventing viral attachment and cell entry [6, 7]. To date, a few investigations have reported the real-world effect of casirivimab-imdevimab as outpatient treatment for mild-to-moderate COVID-19 cases with high-risk aggravations [8–10]. These studies demonstrated that use of casirivimab-imdevimab decreased hospitalization and mortality rates. However, it remains unclear whether casirivimab-imdevimab is also effective for treatment of the SARS-CoV-2 Delta variant.

Starting in March 2020, we have been gathering the medical information of patients with COVID-19, who were hospitalized in 26 medical institutes in Fukushima Prefecture, in our electronic database. A total of 4,568 COVID-19 patients were registered by the end of September 2021. Even in Fukushima Prefecture, the Delta variant was widely spread, and from July to September 2021, the most frequently detected variant of SARS-CoV-2 was the Delta variant [11]. Furthermore, starting in July 2021, casirivimab-imdevimab has been approved in Japan for treating mild-to-moderate COVID-19 patients who are at high-risk of deterioration. Hence, our database was available to evaluate the clinical efficacy of casirivimab-imdevimab against SARS-CoV-2 in the real-world setting.

To the best of our knowledge, the present study is the first real-world retrospective study to evaluate the efficacy of casirivimab-imdevimab for cases of mild-to-moderate COVID-19 caused by the Delta variant of SARS-CoV-2. We compared the clinical outcomes of patients with mild-to-moderate COVID-19 treated with casirivimab-imdevimab therapy to those who underwent alternative treatments.

## 2. Methods

### 2.1. Study design and population

This retrospective cohort study was conducted using an electronic database and enrolled 4,568 COVID-19 patients (as of the end of September 2021) who were admitted to 26 hospitals in Fukushima Prefecture. Those hospitals participated in the web conferences organized by the Department of Pulmonary Medicine, Fukushima Medical University. Among the 4,568 patients, the data of 790 were excluded, because those patients had severe COVID-19, treatment for which casirivimab-imdevimab is not eligible. Among the remaining 3,778 patients, we excluded 2,829 as they were admitted before casirivimab-imdevimab became available on July 24, 2021 in Fukushima prefecture. Finally, we analyzed the data of 949 patients. The subjects in the current study included patients who had been vaccinated against SARS-CoV-2. The clinical characteristics, including comorbidities, examination results, medications, as well as clinical course and outcomes of the subjects, were obtained from the electronic database of each hospital. In the current study, the subjects who received the vaccine twice were categorized as “received vaccination”.

The diagnosis of COVID-19 was made by positive results for SARS-CoV-2 polymerase chain reaction (PCR) on nasopharyngeal swab or saliva samples. Assessment of COVID-19 severity was performed according to the definition issued by the Japanese Ministry of Health, Labor and Welfare: mild, patients without pneumonia or respiratory failure; moderate-1, patients with pneumonia but without respiratory failure; moderate-2, patients with pneumonia and respiratory failure (percutaneous oxygen saturation < 94% on room air) but do not require mechanical ventilation/extracorporeal membrane oxygenation (ECMO); or severe, patients with pneumonia and respiratory failure who require mechanical ventilation/ECMO [12, 13].

Retrospectively, the patients were divided into two groups: (1) those treated with casirivimab-imdevimab; and (2) those not treated with casirivimab-imdevimab (controls). If a patient’s condition deteriorated during the clinical course after the administration of casirivimab-imdevimab, other therapies for COVID-19, including antiviral drugs or immunomodulatory agents (remdesivir, favipiravir, corticosteroid, baricitinib, or tocilizumab), were prescribed and administered at the attending doctor’s discretion.

### 2.2. Patient Eligibility Criteria

The inclusion criteria for treatment with casirivimab-imdevimab were guided by those for the COV-2067 trial [5], and the recommendation of the Japanese Ministry of Health, Labor and Welfare [12]. In particular, patients who were aged ≥ 18 years were eligible for casirivimab-imdevimab treatment if they had symptoms of COVID-19 (e.g., cough, sore throat, fever, and constitutional symptoms), were within 7 days of symptom onset, and had at least one of the following criteria for high risk aggravation: an age of ≥ 50 years, body mass index (BMI) of ≥ 30 kg/m^2,^ use of immunosuppressive medication, immunocompromised status, and presence of hypertension, cardiovascular disease, chronic lung disease, chronic kidney disease (CKD), diabetes mellitus and/or chronic liver disease.

### 2.3. Outcomes of interest

The primary outcomes of interest were any clinical deterioration, need for mechanical ventilation/ECMO, and death after initiation of casirivimab-imdevimab. The secondary outcomes included factors that influenced clinical deterioration.

The definition of clinical deterioration in the present study was a worsened respiratory condition requiring respiratory therapy (use of inhalation oxygen, mechanical ventilation, or ECMO) or the initiation of other medications for COVID-19 (remdesivir, favipiravir, corticosteroid, baricitinib, or tocilizumab) after the first day of hospitalization.

### 2.4. Statistical analyses

Continuous variables are shown as median with interquartile range (IQR) and categorical variables are shown as numbers and percentages. Comparisons between groups for the continuous variables and categorical variables were performed using Mann-Whitney U test and chi-square test, respectively. Age was reclassified as a categorical variable according to the cutoff values obtained from the receiver operating characteristic (ROC) curve of univariate logistic regression analysis for predicting deterioration during hospitalization. Based on the ROC curve and Youden index analysis, we determined that the best cut-off age was 43 (sensitivity 73.6%, specificity 56.4%). Among the comorbidities, those with a prevalence of ≥ 2% were applied to the analyses. The variables that had statistically significant differences between the casirivimab-imdevimab users and non-users were used to identify independent risk factors for predicting deterioration a day or later after admission via multivariate logistic regression analysis. Adjusted odds ratios (OR) with 95% confidence interval (CI) were calculated.

In addition, we compared the risk of exacerbation between the casirivimab-imdevimab users and non-users using the DOAT score. The DOAT score is a simple predictive model that we established and reported in a previous study [14]. It consists of three items: 1) having the comorbidity of diabetes or obesity, 2) being aged ≥ 40 years, and 3) having high body temperature (≥ 38°C). The DOAT score range is 0–3 points, and a high DOAT score (optimal cutoff point is 2) denotes a higher possibility of deterioration in non-elderly COVID-19 patients who do not have respiratory failure on admission.

Furthermore, we used the propensity score technique to match casirivimab-imdevimab users and non-users. The propensity score method selected covariates that may influence casirivimab-imdevimab administration [15]. Logistic regression was performed to estimate the propensity of initiating casirivimab-imdevimab users versus non-user, including 14 confounders that are related to deterioration risk (age, sex, cigarette smoking, high body temperature, percutaneous oxygen saturation, severity, receiving the vaccine, hypertension, obesity, diabetes, dyslipidemia, respiratory diseases, CKD, and collagen vascular disease). We then matched subjects on the logit of the propensity score using a caliper of width equal to 0.2 of the standard deviation of logit of the propensity score [16]. We excluded patients with moderate-2 and severe status from the analyses, because we were unable to match them with control patients. All statistical analyses were performed using JMP 13 (SAS Institute Inc, Cary NC) and EZR (Saitama Medical Center, Jichi Medical University, Saitama, Japan). A two-tailed p-value of < 0.05 was considered statistically significant. Cases with missing data were not excluded from the analyses.

### 2.5. Ethics statement

The need for informed consent was waived because the study is retrospective. This study was approved by the Ethics Committee of Fukushima Medical University (approval number 2020-118, approved on August 3, 2020, updated September 01, 2021).

## 3. Results

### 3.1. Characteristics of participants

The patient selection flowchart is shown in Figure 1. Among the 949 COVID-19 patients enrolled in the current study, 314 were administered casirivimab-imdevimab. The differences in characteristics between the casirivimab-imdevimab users and non-users are demonstrated in Table 1. The casirivimab-imdevimab users were significantly older (P < 0.0001), and had higher body temperature (≥ 38°C) (P < 0.0001) and a higher history of cigarette smoking (P = 0.0068) compared to the non-users. The disease severity was significantly higher in the non-users compared to the casirivimab-imdevimab users (P = 0.0003). Regarding comorbidities, in the casirivimab-imdevimab users, there was a significantly higher prevalence of hypertension (P = 0.0004), obesity (P < 0.0001), and dyslipidemia (P < 0.0001). There were no significant differences in sex, diabetes, chronic respiratory diseases, malignancies, cardiac disease, CKD, and being vaccinated between the two groups. With regard to physical and laboratory examinations, ferritin levels (P = 0.0102) and DOAT score (P < 0.0001) were significantly higher in the casirivimab-imdevimab users than in the non-users. On the other hand, percutaneous oxygen saturation level, white blood cells, neutrophil percentage, lymphocyte percentage, C-reactive protein (CRP), lactate dehydrogenase (LDH), and D-dimer did not significantly differ between the groups.

**Table 1.**
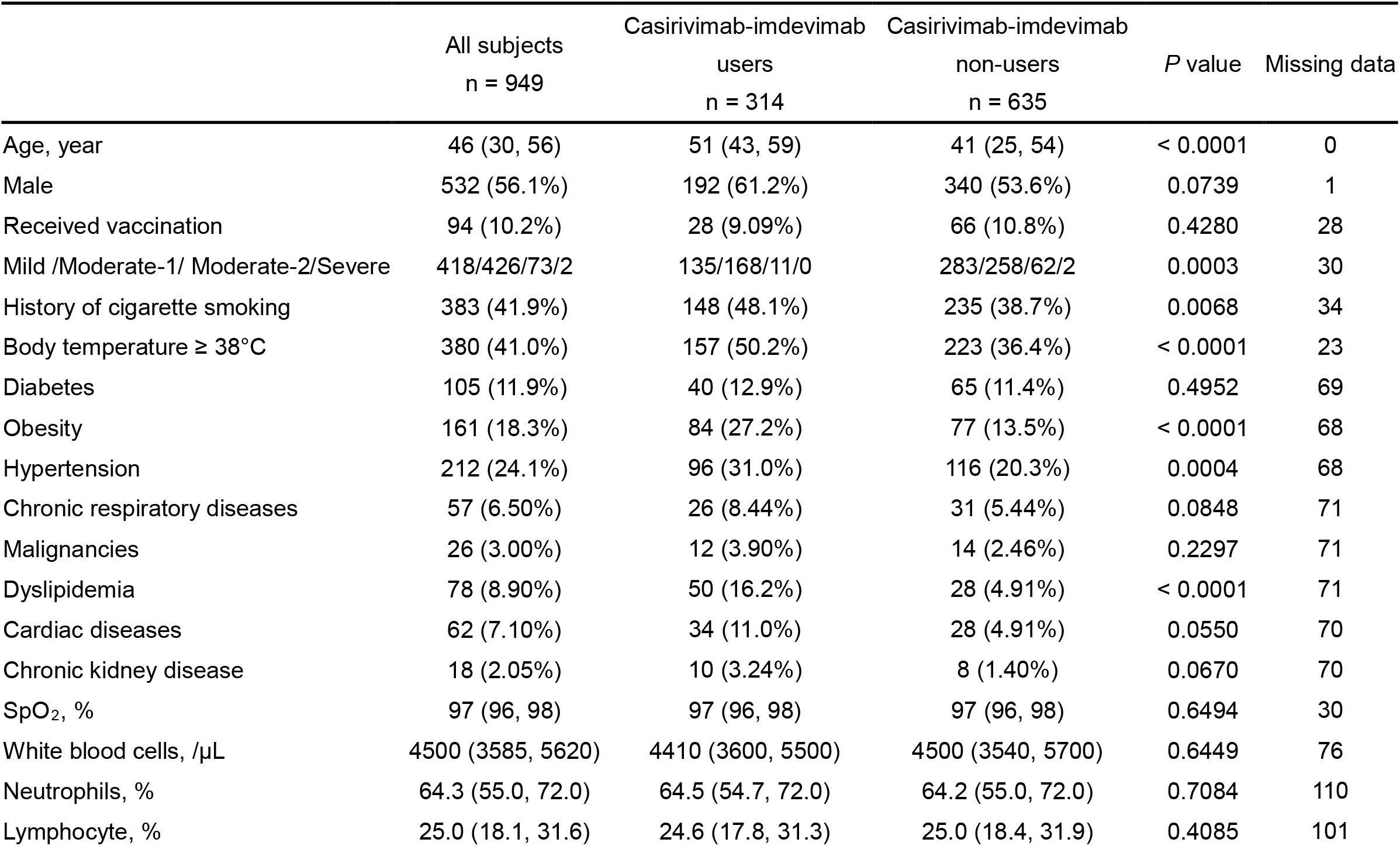
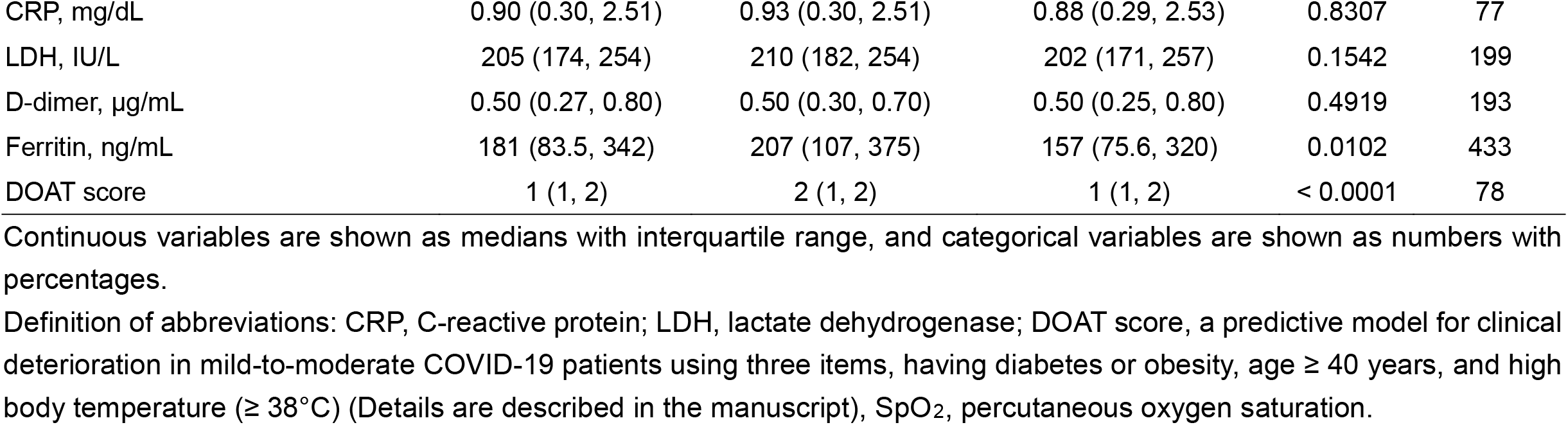
Comparison of baseline clinical characteristics between the casirivimab-imdevimab users and non-users

**Figure 1.**
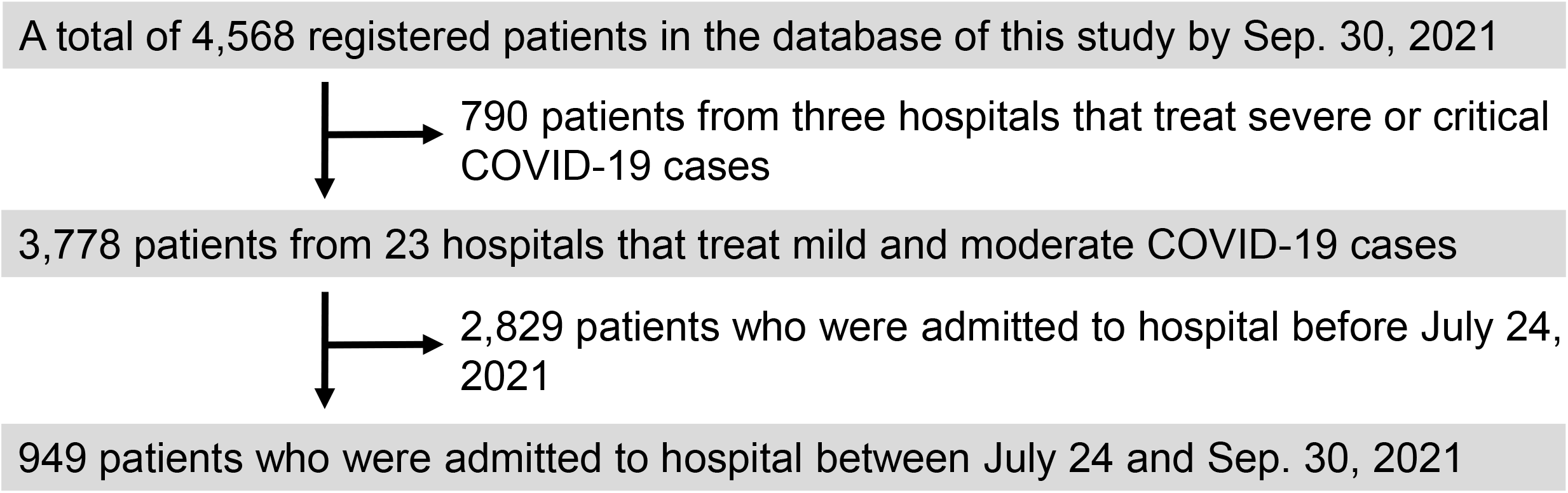
Flowchart of patients’ recruitment in this study Among a total of 4,568 registered COVID-19 patients in our electronic database, 949 were selected for the present study.

### 3.2. Clinical outcomes of treatment with casirivimab-imdevimab

Among all patients analyzed in the present study, 104 experienced deterioration a day after admission or later. The primary endpoints of the rates of clinical deterioration, need for mechanical ventilation/ECMO, and death for the total population were 11.0%, 0.53%, and 0.63%, respectively. One casirivimab-imdevimab user with moderate-1 status died, and there were no significant differences regarding rates of clinical deterioration, need for mechanical ventilation/ECMO and death between the two groups (Table 2).

**Table 2.**
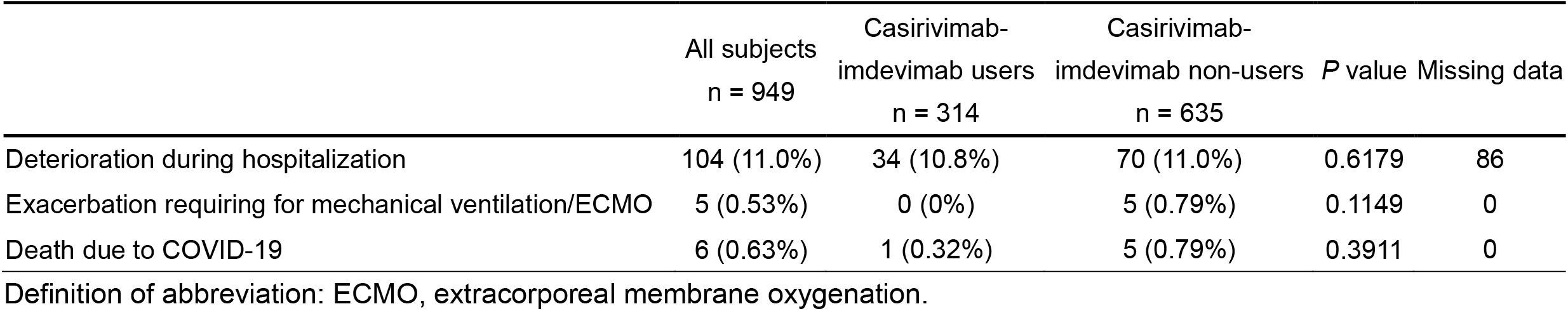
Comparison of the clinical outcomes between the casirivimab-imdevimab users and non-users

|  | All subjects<br>n = 949 | Casirivimab-imdevimab users<br>n = 314 | Casirivimab-imdevimab non-users<br>n = 635 | *P* value | Missing data |
| --- | --- | --- | --- | --- | --- |
| Deterioration during hospitalization | 104 (11.0%) | 34 (10.8%) | 70 (11.0%) | 0.6179 | 86 |
| Exacerbation requiring for mechanical ventilation/ECMO | 5 (0.53%) | 0 (0%) | 5 (0.79%) | 0.1149 | 0 |
| Death due to COVID-19 | 6 (0.63%) | 1 (0.32%) | 5 (0.79%) | 0.3911 | 0 |

Definition of abbreviation: ECMO, extracorporeal membrane oxygenation.

### 3.3. cIndependent risk factors of deterioration after hospitalization

The results of multivariate logistic regression analysis of COVID-19 deterioration during hospitalization are shown in Table 3. According to this analysis, age ≥ 43 years (P = 0.0004), being unvaccinated (P = 0.0002), not receiving casirivimab-imdevimab (P = 0.0023), and dyslipidemia (P = 0.0290) were independent risk factors related to the clinical deterioration of COVID-19 (Table 3).

**Table 3.** Multivariate logistic regression analysis of deterioration among patients with COVID-19 after hospitalization

| | $\beta$ regression coefficient | OR | 95% CI | *P* value |
| --- | --- | --- | --- | --- |
| Age ≥ 43 years-old | 0.466 | 2.543 | 1.491–4.337 | 0.0004 |
| History of cigarette smoking | -0.088 | 0.839 | 0.529–1.328 | 0.4230 |
| Severity, per 1 grade-increase | -0.0158 | 0.984 | 0.678–1.427 | 0.9334 |
| Received vaccination | -0.9565 | 0.147 | 0.042–0.515 | 0.0002 |
| Casirivimab-imdevimab | -0.4006 | 0.448 | 0.263–0.763 | 0.0023 |
| Dyslipidemia | 0.4074 | 2.258 | 1.113–4.582 | 0.0290 |
| Obesity | 0.2572 | 1.672 | 0.963–2.903 | 0.0739 |
| Hypertension | 0.0897 | 1.196 | 0.685–2.087 | 0.5299 |
| Cardiovascular disease | 0.2948 | 1.803 | 0.844–3.853 | 0.1390 |
| Chronic kidney disease | 0.5339 | 2.909 | 0.709–11.929 | 0.1550 |
| Chronic respiratory disease | 0.0129 | 1.026 | 0.413–2.546 | 0.9557 |
| Body temperature ≥ 38°C | 0.0196 | 1.039 | 0.650–1.663 | 0.8704 |

COVID-19 severity grade: mild, subjects without pneumonia or respiratory failure; moderate-1, subjects with pneumonia but without having respiratory failure; moderate-2, subjects with pneumonia and respiratory failure (oxygen saturation < 94% on room air) but who do not require mechanical ventilation/extracorporeal membrane oxygenation (ECMO); and severe, subjects with pneumonia and respiratory failure who require mechanical ventilation/ECMO
Definition of abbreviations: CI, confidence interval; OR, odds ratio.

### 3.4. The clinical deterioration rate between two groups after propensity score matching

Using the propensity score matching method, 222 patients were selected from each group. The baseline clinical characteristics after adjusting for propensity score are summarized in Table 4. There were no significant differences between the two groups, except for the percentage of neutrophils in blood test. The clinical deterioration rate was significantly lower in the casirivimab-imdevimab users compared to the non-users (7.66% vs 14.0%; p = 0.021). None of the 222 patients in both groups required mechanical ventilation/ECMO. One casirivimab-imdevimab user died, but there was no significant difference regarding the death rate between the two groups (Table 5).

**Table 4.** Comparison of baseline clinical characteristics between the casirivimab-imdevimab users and non-users after adjustment with propensity score

|  | Casirivimab-imdevimab users<br>n = 222 | Casirivimab-imdevimab non-users<br>n = 222 | *P* |
| --- | --- | --- | --- |
| Age, year | 50 (39, 57) | 50 (41, 57) | 0.563 |
| Male | 133 (59.9%) | 124 (55.9%) | 0.442 |
| Received vaccination | 22 (9.90%) | 23 (10.4%) | 1.000 |
| Mild/Moderate-1/Moderate-2/Severe | 96/126/0/0 | 93/129/0/0 | 0.848 |
| History of cigarette smoking | 103 (46.4%) | 104 (46.8%) | 0.784 |
| Body temperature $\geq 38^{\circ}\text{C}$ | 96 (43.2%) | 92 (41.4%) | 0.773 |
| Diabetes | 28 (12.6%) | 31 (14.0%) | 0.780 |
| Obesity | 51 (23.0%) | 49 (22.1%) | 0.910 |
| Hypertension | 54 (24.3%) | 57 (25.7%) | 0.827 |
| Chronic respiratory diseases | 16 (7.20%) | 18 (8.1%) | 0.859 |
| Malignancies | 5 (2.30%) | 5 (2.3%) | 1.000 |
| Dyslipidemia | 19 (8.60%) | 16 (7.2%) | 0.725 |
| Cardiac diseases | 17 (7.70%) | 15 (6.8%) | 0.855 |
| Chronic kidney disease | 4 (1.89%) | 3 (1.4%) | 1.000 |
| SpO<sub>2</sub>, % | 97 (96, 98) | 97 (96, 98) | 0.854 |
| White blood cells, /$\mu\text{L}$ | 4400 (3500, 5300) | 4600 (3600, 5700) | 0.150 |
| Neutrophil, % | 63.0 (54.2, 72.0) | 66.3 (57.9, 73.5) | 0.033 |
| Lymphocyte, % | 24.8 (18.3, 31.0) | 23.4 (17.9, 31.1) | 0.412 |
| CRP, mg/dL | 0.70 (0.25, 2.48) | 0.97 (0.38, 2.39) | 0.218 |
| LDH, IU/L | 208 (179, 242) | 208 (173, 260) | 0.529 |
| D-dimer, µg/mL | 0.47 (0.24, 0.66) | 0.47 (0.20, 0.80) | 0.455 |
| Ferritin, ng/mL | 178 (96.0, 348) | 211 (81.4, 377) | 0.696 |

Continuous variables are shown as medians with interquartile range, and categorical variables are shown as numbers with percentages.
Definition of abbreviations: CRP, C-reactive protein; LDH, lactate dehydrogenase; SpO<sub>2</sub>, percutaneous oxygen saturation.

**Table 5.**
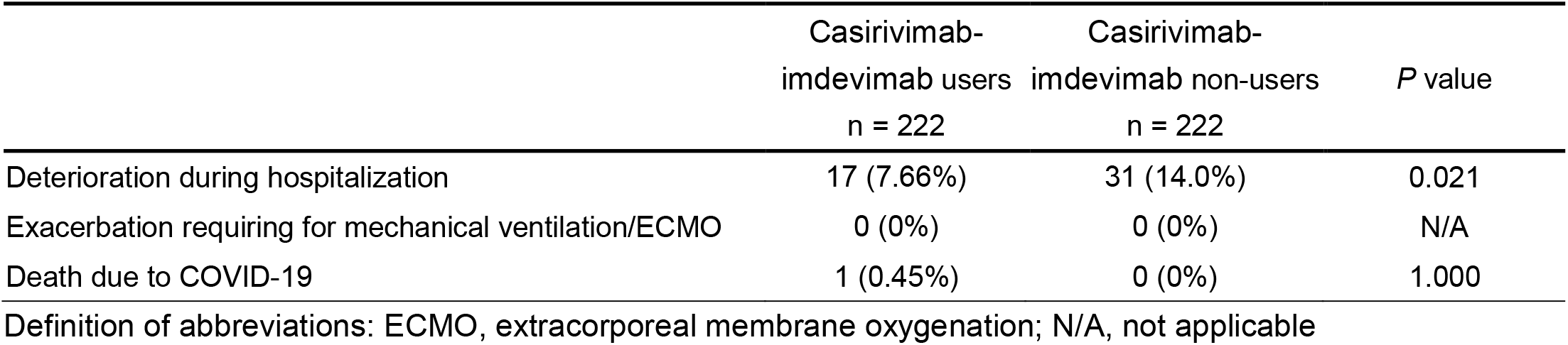
Comparison of the clinical outcomes between the casirivimab-imdevimab users and non-users after adjustment with propensity score

|  | Casirivimab-imdevimab users<br>n = 222 | Casirivimab-imdevimab non-users<br>n = 222 | *P* value |
| --- | --- | --- | --- |
| Deterioration during hospitalization | 17 (7.66%) | 31 (14.0%) | 0.021 |
| Exacerbation requiring for mechanical ventilation/ECMO | 0 (0%) | 0 (0%) | N/A |
| Death due to COVID-19 | 1 (0.45%) | 0 (0%) | 1.000 |

Definition of abbreviations: ECMO, extracorporeal membrane oxygenation; N/A, not applicable

## 4. Discussion

In the present study, we investigated the effectiveness of casirivimab-imdevimab utilizing real-world data from 949 COVID-19 patients who had been admitted to hospitals in Fukushima Prefecture between July 2021 and September 2021 during the Delta variant pandemic. The patients taking casirivimab-imdevimab were older and had more frequent comorbidities such as hypertension, obesity, and dyslipidemia compared to those who were not taking casirivimab-imdevimab. These differences may have been caused by the eligibility criteria of treatment with casirivimab-imdevimab. Thus, it is natural that the DOAT score [14], which is a predictive score of disease progression in COVID-19 patients with mild-to-moderate-1 severity, was higher in the casirivimab-imdevimab users than in the non-users. On the other hand, the disease severity was significantly higher in the non-users. There was no significant difference in clinical outcomes between the two groups in the analysis of data before adjustment for differences in patient characteristics. However, because there were several significant differences in clinical characteristics between the two groups, we performed a multivariate logistic regression analysis that included the confounding factors as explanatory variables. The analysis results demonstrated that not receiving casirivimab-imdevimab was an independent risk factor for deterioration of COVID-19. Furthermore, a propensity score matching analysis for mild-to-moderate patients revealed the effectiveness of casirivimab-imdevimab hidden behind the confounding factors. These results are consistent with a previous study that showed casirivimab-imdevimab treatment to be associated with a significantly lower rate of hospitalization among high-risk patients with mild-to-moderate COVID-19 [9].

In the current study, information on SARS-CoV-2 variants in individual cases was unknown. However, according to a survey performed by the Fukushima Prefectural Institute of Public Health, the proportion of Delta variant cases reached 70% by the end of July 2021 in Fukushima Prefecture, and exceeded 90% after the middle of the following month [11]. Therefore, it is reasonable to consider that almost all cases analyzed in the present study were Delta variant cases. Thus, the results of present study indicate that casirivimab-imdevimab is effective against the SARS-CoV-2 Delta variant.

The Delta variant, which was first detected in India in 2020, has attained global dominance with a higher transmissibility and immune evasion [17]. In vitro and in vivo, casirivimab-imdevimab retains neutralization potency against SARS-CoV-2 variants, including B.1.1.7 (Alpha), B.1.351 (Beta), B.1.617.2 (Delta), B.1.429 (Epsilon), and P.1 (Gamma), and may prevent the selection of resistant variants [18, 19]. According to a previous report by Wilhelm A et al., in which a cell culture-based neutralization assay focusing on the Delta variant was performed, the Delta variant was resistant to imdevimab alone; however, they hypothesized that this might be circumvented by combination treatment with casirivimab [20]. Recently, Kumar et al. reported in their single-center prospective cohort study in India that the use of casirivimab-imdevimab for treating high-risk SARS-CoV-2 patients infected with the Delta variant demonstrated faster improvement of symptoms along with reduced viral loads compared with patients who had been administered remdesivir [21]. Although their results support those of the present study, the details regarding the disease severity and the deterioration rate in the patients taking casirivimab-imdevimab in their study are unknown. Taking our real-world results together with those from Kumar et al., we believe that casirivimab-imdevimab is suggested to be effective for the treatment of the SARS-CoV-2 Delta variant.

The efficacy of SARS-CoV-2 vaccine against the Delta variant was also revealed by multivariate logistic regression analysis in the present study (Table 3). Full vaccination significantly prevented deterioration in the mild-to-moderate COVID-19 patients (OR 0.147, 95% CI 0.042–0.515). Simultaneously, our study showed that casirivimab-imdevimab contributes to the prevention of deterioration (OR 0.448, 95% CI 0.263–0.763), regardless of having received a SARS-CoV-2 vaccine. The effectiveness of SARS-CoV-2 vaccination remains unclear for some variants. The current vaccines have been effective against some variants, such as the B.1.1.7 (Alpha), B.1.351 (Beta), and B.1.617.2 (Delta) variants [22, 23], but have reportedly been less effective against the B.1.621 (Mu) variant [24]. Although further investigation is still required, combination use of SARS-CoV-2 vaccine and casirivimab-imdevimab may synergistically reduce disease progression and mortality of patients with COVID-19 caused by variants.

The strength of the current study is that its results are considered to be generalizable because the population was comprised of inpatients at major hospitals in Fukushima Prefecture that handle COVID-19 inpatient treatment. During the study period, in Fukushima Prefecture, which has a total population of approximately 2 million people, there were 4,155 people infected with COVID-19 [25], and 23% of those patients were registered in the database of the current study.

There are several limitations to the present study. First, this was an observational and retrospective study, and does not show evidence comparable to a randomized clinical trial. Although we carefully adjusted clinical outcomes for various potential cofounding factors, potential bias due to the study design is inevitable. Second, in some subjects, the precise information about the number of days between COVID-19 symptom onset and the administration of casirivimab-imdevimab, and the day of deterioration were not included in our electronic database. Therefore, there may be some influence on the clinical outcomes. Third, the adverse effects of casirivimab-imdevimab were not evaluated in detail. However, according to previous studies, adverse effects were uncommon and mild [5, 10]. Fourth, information regarding which SARS-CoV-2 variant each individual had was not included in the current study. However, as we mentioned above, almost all patients were considered to be infected with the Delta variant during the study period in most of Japan [3], including Fukushima Prefecture [11]. Fifth, regarding the vaccinated patients information on how many days before disease onset they completed SARS-CoV-2 vaccination was not available.

Although the vast majority of healthy individuals respond to the vaccines and obtain full efficacy against SARS-CoV-2 at least 7 days after second vaccination [26], some of our subjects may have been infected with the virus shortly after their second vaccination. The period between the second vaccination and onset of COVID-19 may have influenced some of the results obtained in the present study. However, this effect seems to be minor, because we demonstrated the effectiveness of casirivimab-imdevimab regardless of vaccination in our multivariate logistic regression analysis (Table 3). Sixth, as the study population was almost entirely Japanese, different results may be obtained with different ethnic population. Further study is needed to validate these findings in other ethnic populations.

## 5. Conclusion

This real-world retrospective study of high-risk patients with mild-to-moderate COVID-19 demonstrated a low rate of clinical deterioration after treatment with casirivimab-imdevimab, regardless of whether the patients had received a SARS-CoV-2 vaccine. Aggressive use of casirivimab-imdevimab should be considered to prevent deterioration in high-risk patients with mild-to-moderate COVID-19.

## Data Availability

All data produced in the present work are contained in the manuscript.

## Abbreviations

BMI: body mass index
CI: confidence interval
CKD: chronic kidney disease
COVID-19: coronavirus disease 2019
CRP: C-reactive protein
ECMO: extracorporeal membrane oxygenation
IQR: interquartile range
LDH: lactate dehydrogenase
OR: odds ratio
PCR: polymerase chain reaction
SARS-CoV-2: severe acute respiratory syndrome coronavirus 2

## Acknowledgements

We thank the Scientific English Editing Section of Fukushima Medical University for their fruitful discussion and linguistic assistance in proofreading the manuscript.

## Notes

**Conflict of Interest** Yoko Shibata and Hiroyuki Minemura received lecture fees and research grants from Chugai Pharmaceutical Co., Ltd. The other authors report no conflicts of interest related to this study.

### Competing Interest Statement

Yoko Shibata and Hiroyuki Minemura received lecture fees and research grants from Chugai Pharmaceutical Co., Ltd. The other authors report no conflicts of interest related to this study.

### Funding Statement

This study did not receive any funding.

